# Mendelian randomization analysis identifies blood tyrosine levels as a biomarker of non-alcoholic fatty liver disease

**DOI:** 10.1101/2021.11.26.21266879

**Authors:** Émilie Gobeil, Ina Maltais-Payette, Nele Taba, Francis Brière, Nooshin Ghodsian, Erik Abner, Jérôme Bourgault, Éloi Gagnon, Hasanga D. Manikpurage, Christian Couture, Patricia L. Mitchell, Patrick Mathieu, François Julien, Jacques Corbeil, Marie-Claude Vohl, Sébastien Thériault, Tõnu Esko, André Tchernof, Benoit J. Arsenault

**Affiliations:** Centre de recherche de l’Institut universitaire de cardiologie et de pneumologie de Québec, Québec (QC), Canada; Estonian Genome Center, Institute of Genomics, University of Tartu, Tartu, Riia 23b, 51010, Estonia; Institute of Molecular and Cell Biology, University of Tartu, Tartu, Riia 23, 51010, Estonia; Department of Surgery, Faculty of Medicine, Université Laval, Québec (QC), Canada; Department of molecular medicine, Faculty of Medicine, Université Laval, Québec (QC), Canada; Centre de recherche du CHU de Québec, Québec (QC), Canada; Centre NUTRISS, Institut sur la nutrition et les aliments fonctionnels, Université Laval, Québec (QC), Canada; School of Nutrition, Université Laval, Québec (QC), Canada; Department of Molecular Biology, Medical Biochemistry and Pathology, Faculty of Medicine, Université Laval, Québec (QC), Canada; Department of Medicine, Faculty of Medicine, Université Laval, Québec (QC), Canada

**Author notes:** <u>Address for correspondence</u> Benoit Arsenault, PhD, Centre de recherche de l’Institut universitaire de cardiologie et de pneumologie de Québec – Université Laval, Y-3106, Pavillon Marguerite D’Youville, 2725 chemin Ste-Foy, Québec (QC), Canada G1V 4G5.

## Abstract

Non-alcoholic fatty liver disease (NAFLD) is a complex cardiometabolic disease associated with premature mortality. The diagnosis of NAFLD is challenging and the identification of biomarkers causally influenced by NAFLD may be clinically useful. We aimed at identifying blood metabolites causally impacted by NAFLD using two-sample Mendelian randomization (MR) with validation in a population-based biobank and a cohort of patients undergoing bariatric surgery. Our instrument for genetically-predicted NAFLD (the study exposure) included all independent genetic variants (n=7 SNPs) from a recent genome-wide association study on NAFLD. The study outcomes included 123 blood lipids, lipoproteins and metabolites measured in 24,925 individuals from 10 European cohorts. After correction for multiple testing, we identified a positive effect of NAFLD on plasma tyrosine levels but not on other metabolites. The association between NAFLD and tyrosine levels was consistent across MR methods and robust to outliers and pleiotropy. In observational analyses performed in the Estonian Biobank (10,809 individuals including 359 patients with NAFLD), after multivariable adjustment, tyrosine levels were positively associated with the presence of NAFLD (odds ratio per 1-SD increment = 1.23 (95% confidence interval = 1.12-1.36, p = 2.19e-05). In a sample of 138 patients undergoing bariatric surgery, compared to patients without NAFLD, blood tyrosine levels were higher in those with NAFLD, but were comparable among patients with or without non-alcoholic steatohepatitis. This analysis revealed a potentially causal effect of NAFLD on blood tyrosine levels, suggesting that blood tyrosine levels may represent a new biomarker of NAFLD.

**Graphical abstract:** 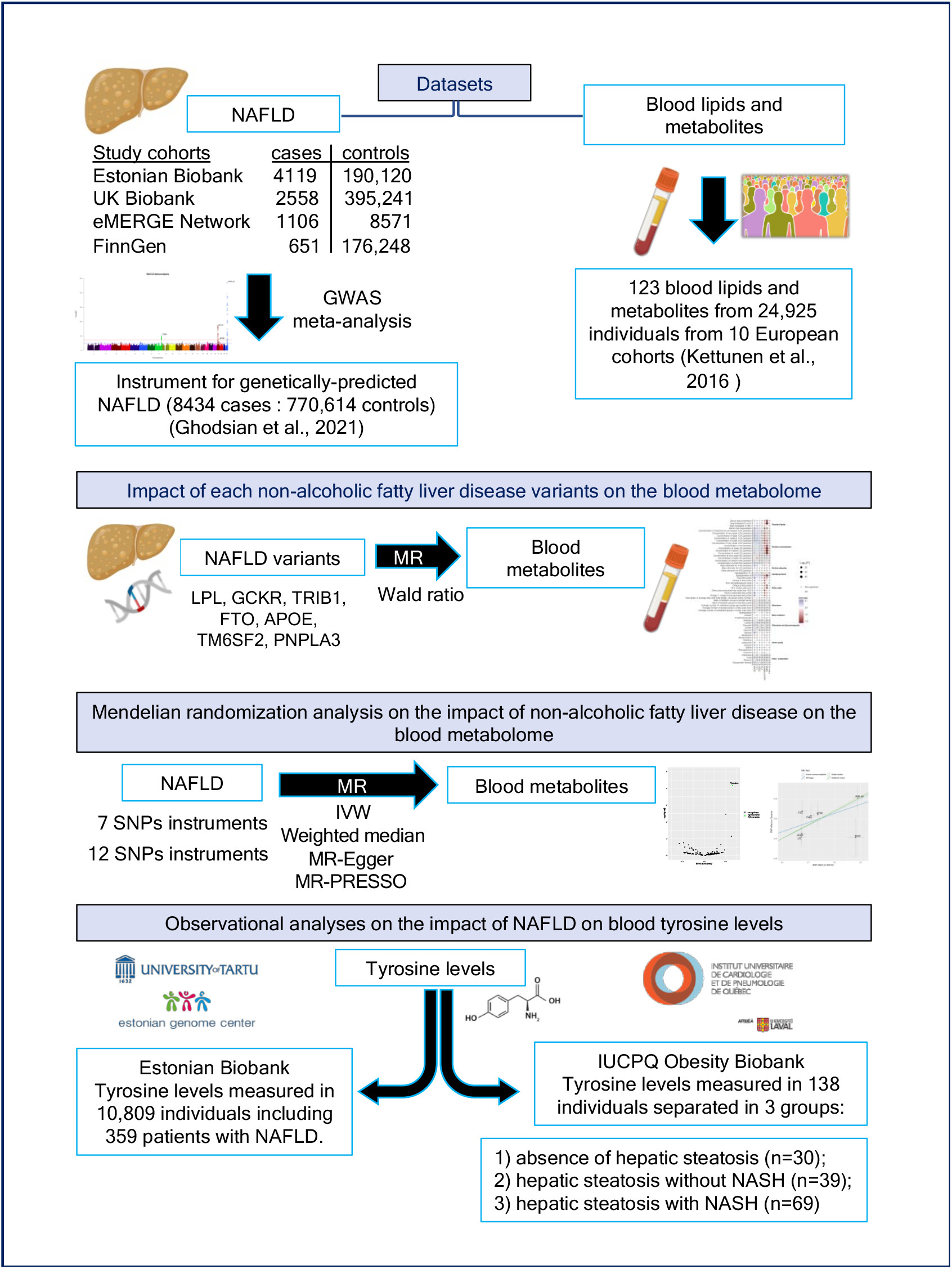

## Introduction

Non-alcoholic fatty liver disease (NAFLD) is the most common liver disease with an estimated prevalence of one in four adults in most Western countries^1^. NAFLD is a progressive disease initiated by the accumulation of lipid droplets within hepatocytes which can lead to inflammation, cell death and to more advanced stages such as non-alcoholic steatohepatitis (NASH) (with or without fibrosis), cirrhosis and liver cancer. Cardiovascular diseases are the leading cause of death in patients with NAFLD^2^. This condition is also associated with other comorbidities such as type 2 diabetes, chronic kidney disease and gastrointestinal neoplasms^3-5^. There is currently no pharmacological treatment available specifically for the treatment of NAFLD.

According to the National Institutes of Health U.S. National Library of Medicine, there are currently more than 300 ongoing randomized clinical trials (RCTs) enrolling patients with NAFLD. Such RCTs are challenging because NAFLD “diagnosis” often requires invasive methods and/or imaging approaches, which are clinically burdensome and cost-prohibitive, especially since NAFLD has reached epidemic proportions in developing countries that may not have the clinical, financial, and infrastructural resources to identify and adequately treat patients with NAFLD. For example, liver biopsy is not only invasive and expensive but is also prone to sampling error.^6^ Affordable and easily obtainable tests are required to identify NAFLD patients who may benefit from therapies under investigation. Blood biomarkers of NAFLD that are not modulated by secondary non-causal pathways, may be promising candidates for the identification of at-risk individuals and to develop tailored therapy for NAFLD.

Mendelian randomization (MR) is a modern epidemiology investigation technique that is increasingly used to explore whether risk factors associated with disease traits reflect true causal associations or not.^7^ Akin to a RCT, MR takes advantage of the random allocation of genetic variation at conception to explore whether human traits that are at least in part under genetic control are associated with diseases. MR has also been used to determine whether a genetic susceptibility to certain diseases influences other biological traits such as the blood metabolome, thereby identifying early biomarkers of disease-related traits.^8,9^

High-quality MR studies rely on the availability of standardized effect sizes of the major genetic variants associated with a trait of interest when that trait is used as an exposure and on the availability of genome-wide association study (GWAS) summary statistics when that trait is used as an outcome. In a recent study^10^, we performed a GWAS meta-analysis of NAFLD in four cohorts totaling 8434 NAFLD cases and 770,180 controls. This analysis identified genetic variants at the *GCKR, LPL, TRIB1, FTO, TM6SF2, APOE* and *PNPLA3* as NAFLD susceptibility loci. In this study, we used a combination of observational and two-sample MR study designs to identify blood metabolites that may be causally influenced by the presence of NAFLD.

## Results

### Impact of non-alcoholic fatty liver disease variants on the blood metabolome

We first investigated the impact of each genetic variant associated with NAFLD (at the *GCKR, LPL, TRIB1, FTO, TM6SF2, APOE* and *PNPLA3* loci) on the blood metabolome using GWAS summary statistics of 123 blood lipids, lipoproteins and metabolites measured in 24,925 individuals from 10 European cohorts, as described by Kettunen et al.^11^ The genetic variants selected for this analysis and their effect size on NAFLD are presented in Supplementary Table 1. Figure 1 presents the impact of each NAFLD variant on selected markers of the blood metabolome. This analysis showed that the lead NAFLD variant at the *GCKR* locus was associated with several blood metabolites such as higher lipoprotein/lipid levels, branched chain amino acids and other amino acids as well as glycolysis and gluconeogenesis metabolites. The lead NAFLD variants at the *LPL* and *TRIB1* loci were also associated with higher lipoprotein/lipid levels, while the lead NAFLD variant at the *MAU2/TM6SF2* and *APOE* loci were associated with lower lipoprotein/lipid levels. The NAFLD associated variants at the *FTO* and *PNPLA3* loci did not appear to have a major influence on the blood metabolome.

**Figure 1.**
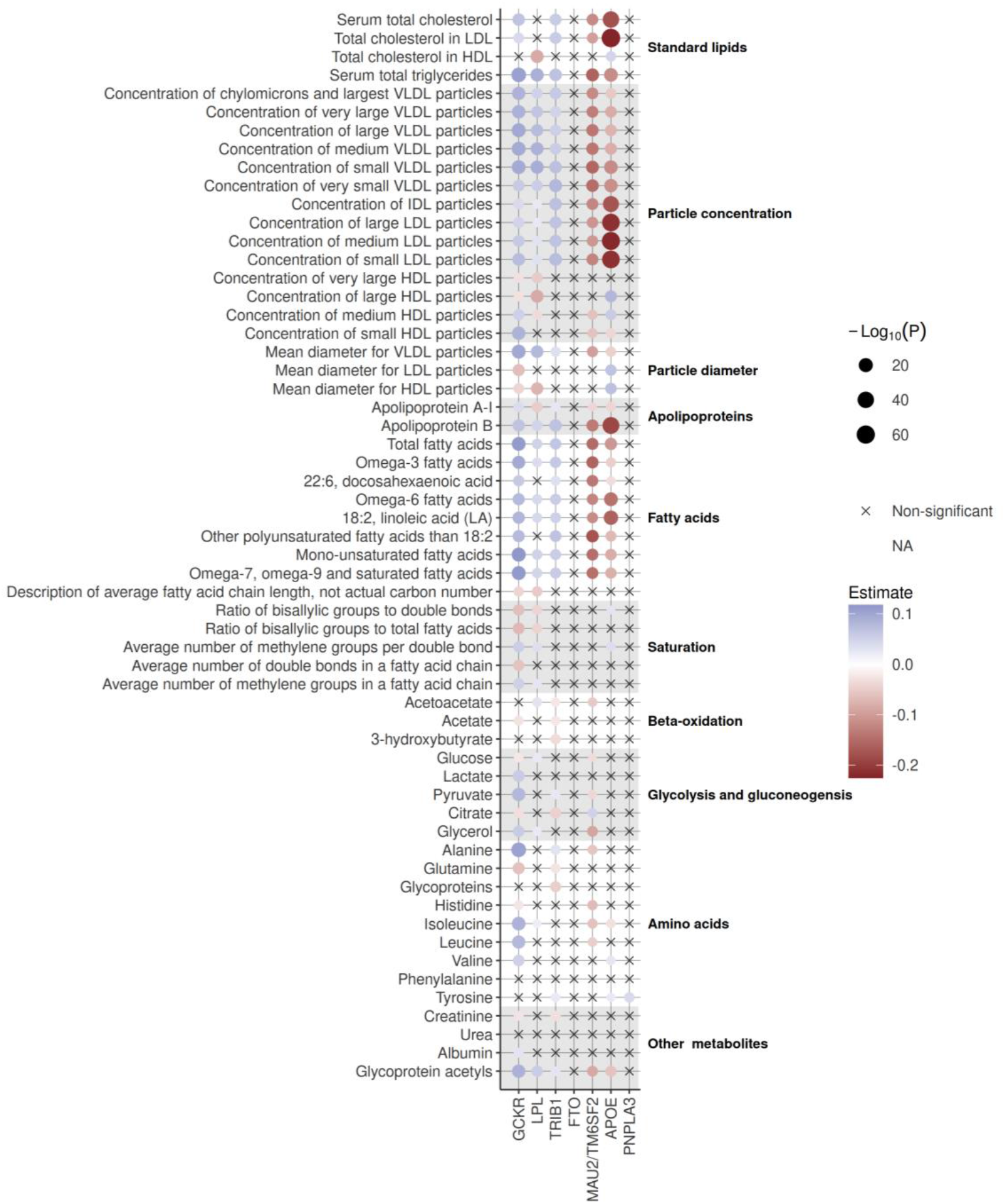
Causal impact of non-alcoholic fatty liver disease (NAFLD) variants on the blood metabolome. Balloon plot reflecting the effect of each of the seven NAFLD variants on 123 blood metabolites using inverse-variance weighted Mendelian randomization. Since an important proportion of metabolites were highly correlated, only groups of metabolites with significant associations after Bonferroni correction are displayed (n=58 metabolites).

### Mendelian randomization analysis on the impact of non-alcoholic fatty liver disease on the blood metabolome

Next, we explored the potentially causal effect of genetically-predicted NAFLD on blood metabolites, using genetic instruments in instrumental-variable analysis implemented via two-sample MR. For performing two-sample MR, we used the summary statistics of two GWAS: NAFLD (exposure) and 123 metabolites (outcome). Using inverse-variance weighted (IVW)-MR, we found that genetically-predicted NAFLD was robustly associated with higher levels of tyrosine (p= 6.75E-05) after correction for false-discovery rate with the Benjamini-Hochberg method (pFDR<4.06E-04 [0.05/123 metabolites]) (Figure 2). We also found an association between NAFLD and the tyrosine precursor, phenylalanine (p=0.0035, Supplementary Table 2), although this association did not pass the FDR-corrected statistical significance threshold. The association between NAFLD and tyrosine levels was consistent across MR methods and robust to outliers and pleiotropy (Table 1 and Figure 3). Because there was sample overlap between the exposure (genetically-predicted NAFLD) and outcomes (blood metabolites), with the Estonian Biobank contributing to both datasets, we performed another NAFLD GWAS meta-analysis, which excluded the Estonian Biobank participants. Genetically predicted NAFLD was still associated with tyrosine (beta [SE] = 0.085 [0.026], p=9.37E-04) using IVW-MR. One of the key assumptions of MR is that genetic variants used as a proxy of the exposure influence the outcome via their effect on the exposure and not via other related traits (horizontal pleiotropy)^12,13^. Our original NAFLD GWAS identified 7 NAFLD susceptibility loci^14^. Some of these loci were identified after leveraging prior effect sizes of NAFLD-related traits such as body mass index and triglycerides, which might increase the chance of finding associations that may be influenced by NAFLD related traits and not by NAFLD per se. We therefore investigated the impact of genetically-predicted NAFLD on blood levels of tyrosine using 12 independent NAFLD SNPs from our original GWAS. For that purpose, we used NAFLD-associated SNPs with p-value for association <5e-06 and r2<0.1). Multiple MR methods were used to investigate the association of genetically-predicted NAFLD with tyrosine levels. The independence of genetic instruments was ensured by obtaining the LD matrix using the European 1000-genome LD reference panel (Supplementary Table 3). Results presented in Table 1 suggest that genetically-predicted NAFLD was strongly associated with tyrosine levels using this other genetic instrument that might be less susceptible to horizontal pleiotropy.

**Table 1.** Association of genetically-predicted non-alcoholic fatty liver disease with blood tyrosine levels across multiple Mendelian randomization methods.

| Metabolite | Inverse-variance weighted |  |  |  | Simple median |  |  | Weighted median |  |  | MR-Egger |  | MR PRESSO |
| --- | --- | --- | --- | --- | --- | --- | --- | --- | --- | --- | --- | --- | --- |
|  | N<br>SNPs | Beta | SE | P-value | Beta | SE | P-value | Beta | SE | P-value | Intercept | intercept<br>P-value | outlier test<br>P-value |
| Tyrosine | 7 | 0.131 | 0.033 | 6.75E-05 | 0.142 | 0.045 | 0.002 | 0.137 | 0.038 | 2.82E-04 | 0.010 | 0.515 | 0.334 |
| Tyrosine | 12 | 0.104 | 0.027 | 1.47E-04 | 0.042 | 0.040 | 0.297 | 0.103 | 0.036 | 3.88E-03 | -0.002 | 0.759 | 0.293 |

The effect of genetically-predicted non-alcoholic fatty liver disease on tyrosine levels using two genetic instruments (one with 7 genome-wide significant variants and one with the 12 independent NAFLD variants with p-value for NAFLD associations <5E-06 are presented.

**Figure 2.**
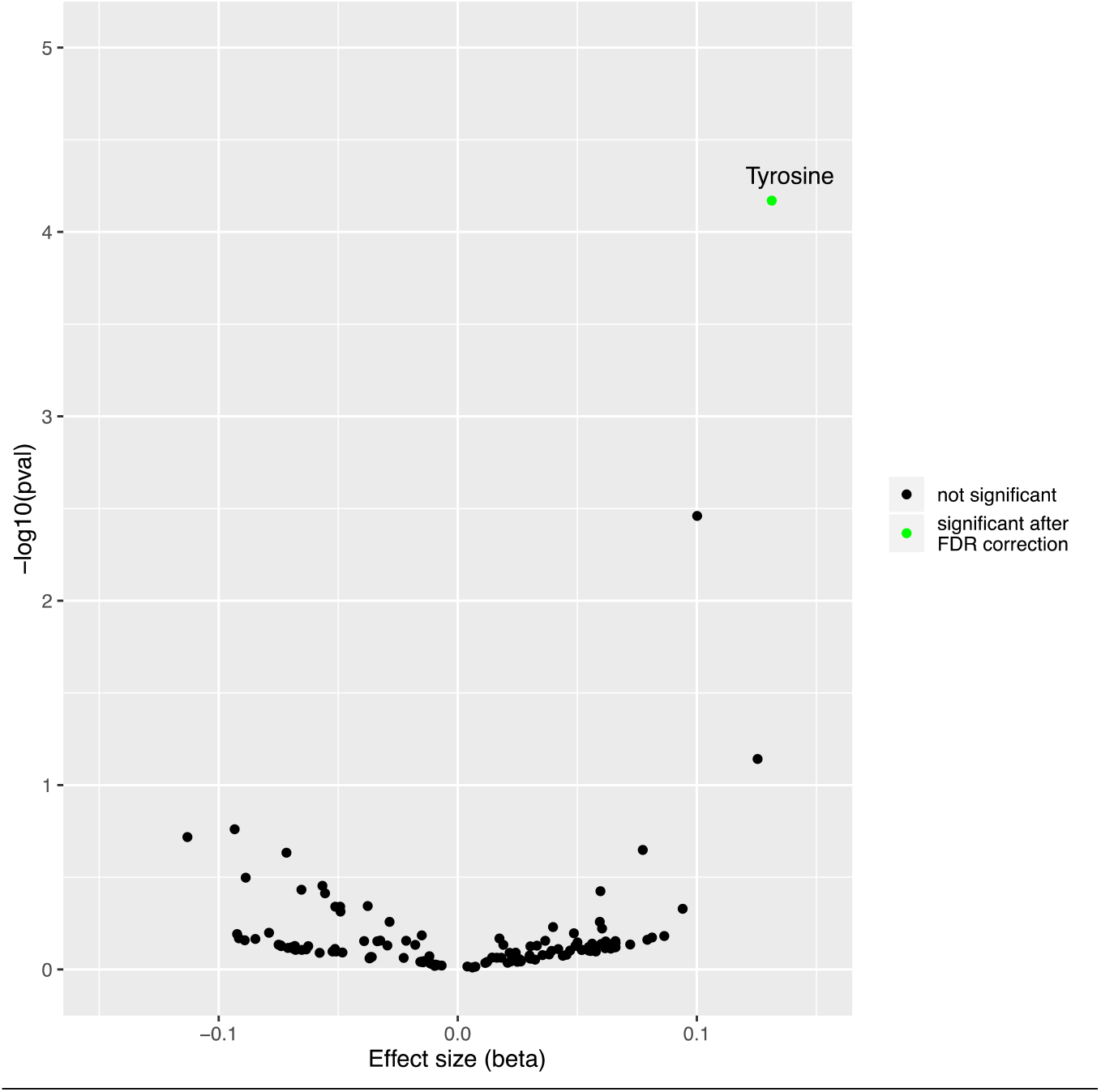
Causal impact of genetically predicted non-alcoholic fatty liver disease (NAFLD) on the blood metabolome. Volcano plot depicting the effect of genetically-predicted NAFLD on blood metabolites (n=123) using inverse-variance weighted Mendelian randomization. Each dot represents a different metabolite, and the green dot represents the metabolite significantly influenced by the presence of NAFLD (tyrosine) following correction for false discovery rate (pFDR<0.05/123 metabolites).

**Figure 3.**
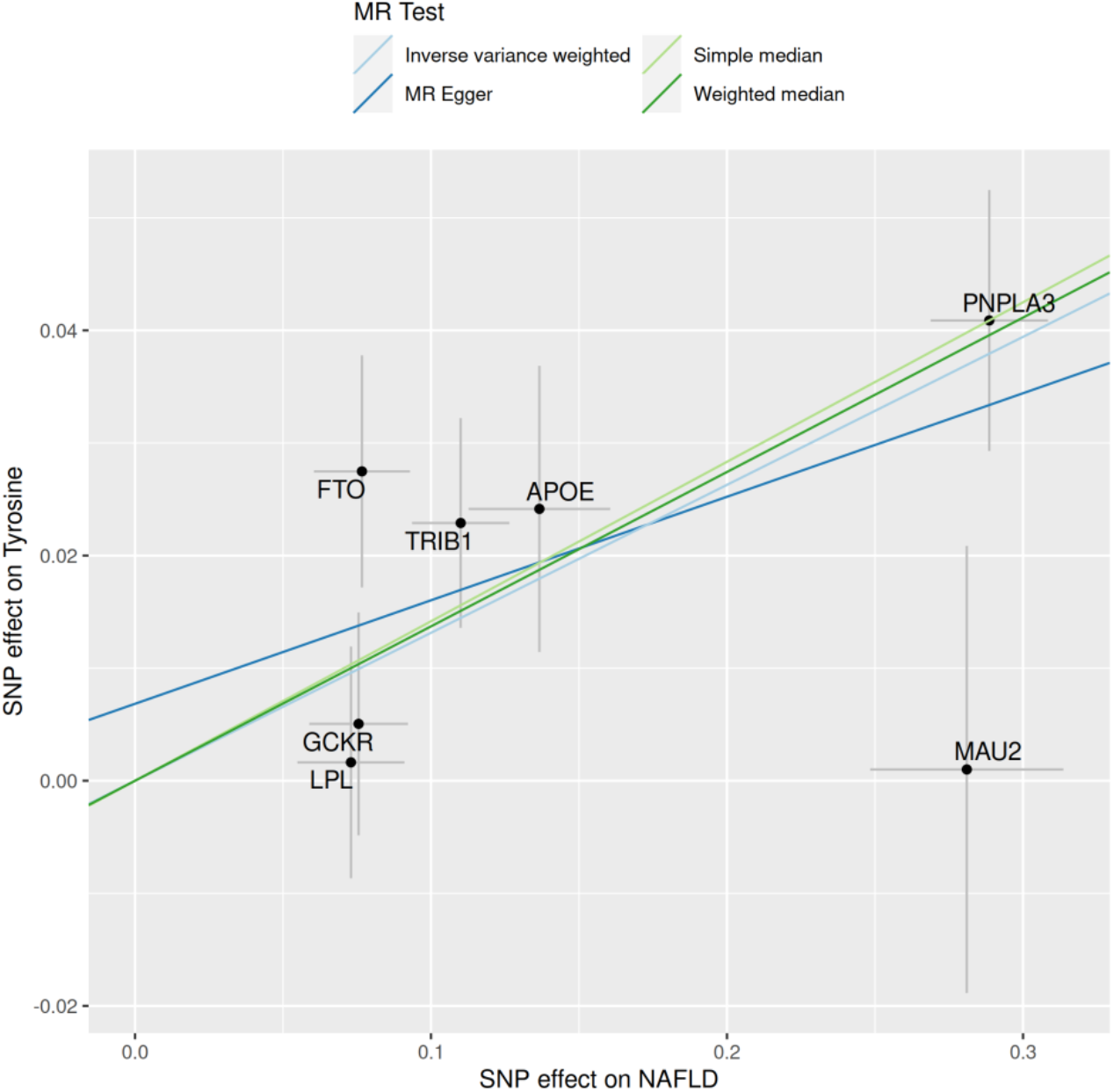
A Mendelian randomization study of genetically-predicted non-alcoholic fatty liver disease and plasma levels of tyrosine. Scatter plot showing the estimated effect sizes of each of the 7 genetic loci associated with NAFLD on NAFLD and blood tyrosine levels and the regression slopes of four MR methods (inverse-variance weighted, simple median, weighted median and MR-Egger).

### Observational analysis on the impact of non-alcoholic fatty liver disease on blood tyrosine levels in the Estonian Biobank

We next investigated whether the presence of NAFLD was associated with higher plasma levels of tyrosine in the Estonian Biobank. Tyrosine levels were measured in 10,809 individuals including 359 patients with NAFLD (obtained from EHR). Table 2 presents the association between tyrosine levels per one-standard deviation increment and NAFLD before and after multivariable adjustment. After adjusting for age, sex, smoking, education, and BMI, tyrosine levels were positively associated with the presence of NAFLD (odds ratio per 1-SD increment = 1.23 [95% confidence interval = 1.12-1.36, p = 2.19e-05]). Altogether, these results provide validation from an observational study of the association of NAFLD with plasma tyrosine levels.

**Table 2.**
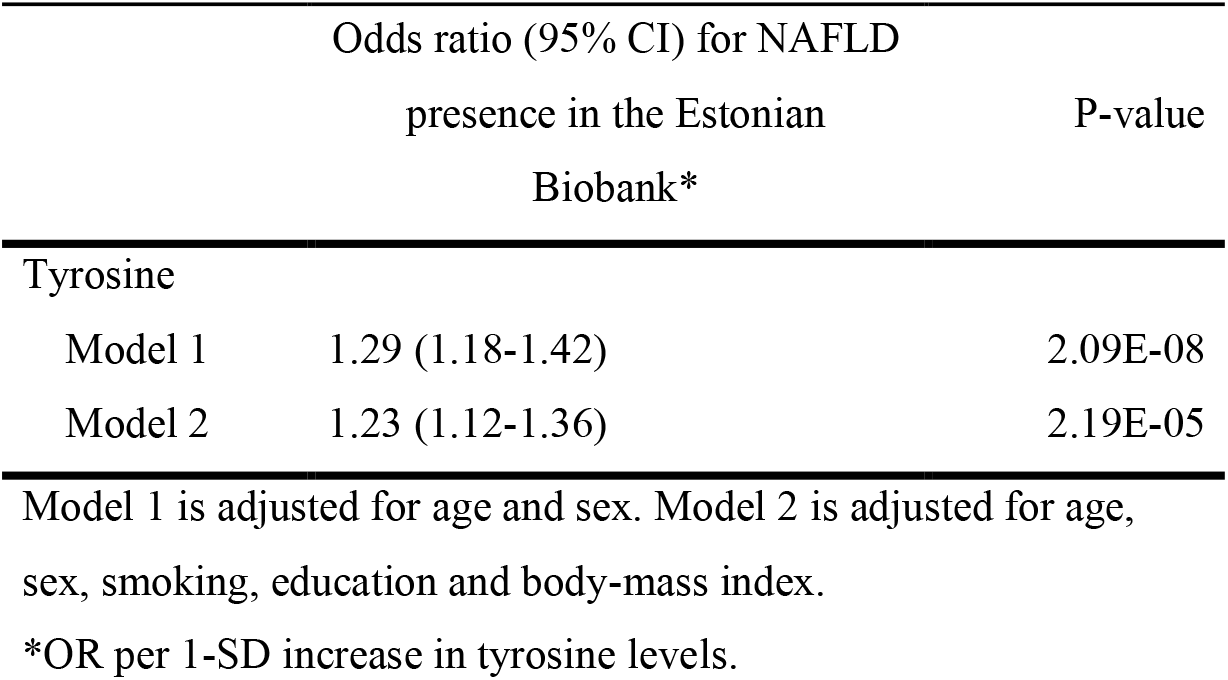
Association of plasma tyrosine levels (SD) with the presence of non-alcoholic fatty liver disease in the Estonian Biobank.

### Impact of non-alcoholic steatohepatitis on tyrosine levels in patients undergoing bariatric surgery

Although an elevated body weight is an important risk factor for NAFLD, the presence and severity of NAFLD is very heterogeneous among patients with obesity. Whether blood-based biomarkers of NAFLD such as tyrosine levels could help identify and stratify patients with NAFLD/NASH is unknown. We therefore investigated whether tyrosine levels were associated with the presence of NAFLD with or without histologically confirmed NASH among 138 participants of the IUCPQ Obesity Biobank. Supplementary Table 4 presents the clinical information at the time of surgery for patients of each group (healthy liver, NAFLD without or with NASH). Compared to patients without NAFLD, blood tyrosine levels were higher in those with NAFLD (N=108). However, among patients with NAFLD, tyrosine levels were comparable among patients with or without NASH (Figure 4). These results suggest that plasma tyrosine levels could be useful to identify patients with NAFLD among patients with obesity, but it may not identify patients with a more advanced stage of NAFLD such as NASH.

**Figure 4.**
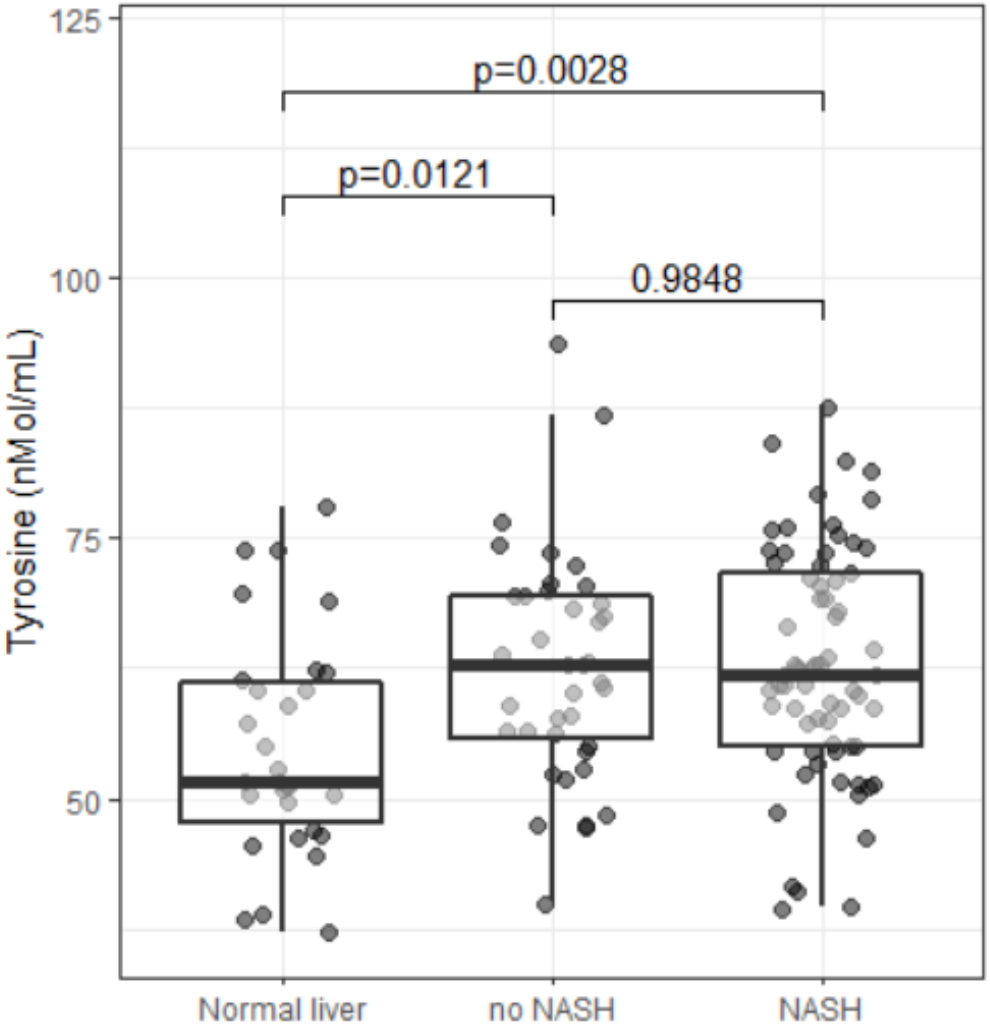
Observational analysis of the impact of NAFLD and NASH on tyrosine levels in the IUCPQ Obesity Biobank. Box plot representing the dispersion and the median values of tyrosine levels between the three groups (without NAFLD= 54.9 ± 10.7 nmol/mL; with NAFLD-without NASH= 62.5 ±10.9 nmol/mL; with NAFLD and NASH = 62.9 ±11.0 nmol/mL). P-values are from Tukey HSD test.

## Discussion

We established a MR framework aimed at identifying novel biomarkers of NAFLD. Results of this analysis suggest that genetically-predicted NAFLD may not be causally linked with metabolites associated with triglyceride-rich lipoprotein metabolism, glucose-insulin homeostasis, or branched-chain amino acid levels. However, this MR analysis revealed an effect of NAFLD on blood tyrosine levels, which may represent a new clinical biomarker of NAFLD. We also reported that patients with higher blood levels of tyrosine had a higher prevalence of NAFLD in the Estonian Biobank and that among patients with obesity, blood tyrosine levels were higher in patients with NAFLD. Results, from the two observational studies with different clinical settings (i.e. a population-based biobank and a bariatric surgery cohort), provide validation to our initial findings obtained with MR.

Several observational studies have suggested that liver fat accumulation or NAFLD negatively impacts triglyceride-rich lipoprotein metabolism, glucose-insulin homeostasis as well as branched-chain amino acid levels.^15-19^ Sliz et al.^20^ also documented the individual impact of 4 variants (at the *PNPLA3, TM6SF2, GCKR* and *LYPLAL1* loci) on the blood metabolome and found inconsistent associations. Here, we used a similar approach to investigate the effect of the top NAFLD variants (including 3 already investigated by Sliz et al.). This analysis confirmed that the lead NAFLD variant at the *GCKR* locus is linked with several blood metabolites levels such as higher lipoprotein/lipid, branched chain amino acids and other amino acids as well as glycolysis and gluconeogenesis metabolites. Variation at the *LPL* and *TRIB1* loci were also associated with higher lipoprotein/lipid levels. On the other hand, variation at the *APOE* and *MAU2/TM6SF2* locus was associated with lower lipoprotein/lipid levels. We investigated whether the presence of NAFLD impacted lipoprotein levels and metabolites of these pathways to identify early biomarkers of NAFLD using MR. This analysis did not find evidence of a causal association of NAFLD with triglyceride-rich lipoprotein metabolism, which is expected since some variants were associated with higher lipid levels while other variants were associated with lower lipid levels. NAFLD was not however associated with glucose-insulin homeostasis markers or branched-chain amino acids. We did however find a significant impact of NAFLD on tyrosine and, to a lesser extent, its metabolic precursor phenylalanine. Although the impact of NAFLD on tyrosine metabolism has been reported decades ago^21^, our analysis adds to this body of evidence by suggesting that the impact of NAFLD on tyrosine metabolism might be a direct consequence of NAFLD, and that this association might not be driven by secondary causes of NAFLD.

Previous studies have investigated the link between excess adiposity and its metabolic consequences such as NAFLD and amino acid levels such as tyrosine. A study by Kimberley et al. provided evidence that tyrosine was positively related to waist circumference (WC) and body mass index (BMI) in 997 participants of the Framingham cohort^22^. Another study performed in a cohort of 38 participants in an outpatient clinic (separated in four groups: 10 patients with normal glucose tolerance and NAFLD, 10 patients with T2D and NAFLD, 8 patients with T2D and no liver disease and a group of 10 controls) identified several non-branched-chain amino acids, including tyrosine, that may be increased in patients with NAFLD without T2D^23^. In two other metabolomic studies, Boulet et al. and Brennan et al. showed that tyrosine was positively related to many adiposity indices including abdominal fat cell size and adipose tissue depots^24,25^. Boulet et al. tested the association between levels of 138 metabolites detectable in plasma and adiposity measurements in 59 healthy women. Concentrations of tyrosine were positively associated with visceral adipose tissue area, subcutaneous adipose tissue area and significantly associated with the mean adipocyte diameter in both fat compartments. In a study of 103 middle-aged patients with abdominal obesity Brennan et al. reported significant associations between tyrosine levels and abdominal adipose tissue and between visceral adipose tissue accumulation and phenylalanine, the precursor of tyrosine. Whether these association could be explained by the fact that NAFLD causes elevations in tyrosine levels, which in turn influences the fate of adipocytes needs to be further explored. Furthermore, these studies suggest a disruption of the hepatic amino acid metabolism in the setting of NAFLD, but the mechanisms underlying the relationship between amino acids imbalance is poorly understood. Winther-Sørensen et al. recently reported that with hepatic steatosis had impaired clearance of amino acids^26^. However, tyrosine was not considered in this investigation. In another study, individuals with NAFLD were also characterized by higher gene expression of metabolic enzymes that may influence amino acids release in the circulation^27^.

Our study has limitations. For instance, although we used of the largest NAFLD dataset available to date and have excluded secondary causes of NAFLD whenever possible, an EHR-based diagnosis of complex diseases such as NAFLD might be prone to misclassification of cases and controls. The prevalence of NAFLD was also not available in some of the cohorts used to document the impact of NAFLD on the blood metabolome (24,925 individuals from 10 European cohorts). Studies documenting the impact of NAFLD resolution on tyrosine levels could also consolidate the causal effect of NAFLD on the blood metabolome.

In conclusion, our study identified a blood metabolite, the amino acid tyrosine, that may be causally influenced by the presence of NAFLD. These findings shed light on the metabolic consequences of NAFLD but also identify potential early biomarkers of NAFLD that could be used to identify patients who may benefit from therapies targeting NAFLD and/or for risk stratification in this population. Additional studies will be required to determine whether our findings could be helpful to optimizing NAFLD risk prediction as well as patient recruitment for trials aiming at preventing and/or treating NAFLD.

## Methods

### Impact of NAFLD on the blood metabolome

To perform the MR analysis, we combined information of two publicly available GWAS summary statistics in a two-sample MR setting. Genetic association estimates for NAFLD (exposure) were obtained from our recently published GWAS^14^ (8434 cases and 770,180 controls) of European ancestry from four cohorts. Briefly, we performed a fixed effect GWAS meta-analysis of the Estonian Biobank, the UK Biobank, The Electronic Medical Records and Genomics (eMERGE)^28^ network, and FinnGen with the *METAL* package^29^. NAFLD cases were obtained by electronic health record codes or hospital records. For the logistic regression analysis, we performed an adjustment for age, sex, genotyping site and the first three ancestries based principal components. For MR analysis with the instrument for NAFLD of 7 variants, we selected lead SNPs, associated with NAFLD through risk factor informed GWAS, for each risk locus (at the *GCKR, LPL, TRIB1, FTO, TM6SF2, APOE* and *PNPLA3* loci) (Supplementary Table 1). For the selection of the 12 SNPs instrument of genetically-predicted NAFLD, SNPs with a p≤5e-06 were kept and we ensured the independence of genetic instruments by clumping all neighbouring SNPs in a 10 Mb window with a linkage disequilibrium r2<0.1 using the European 1000-genome LD reference panel (Supplementary Table 3). We used GWAS summary statistics from the study of Kettunen et al.^11^ to define our study outcomes. In this study, 123 blood lipids and metabolites were measured in 24,925 individuals from 10 European cohorts using high-throughput nuclear magnetic resonance spectroscopy. Metabolites measured using this platform represent a broad molecular signature of systemic metabolism and include metabolites from multiple metabolic pathways (mostly lipoprotein lipids and subclasses, fatty acids as well as amino acids, glycolysis precursors, etc.). The estimates of the metabolites were normalized and reported on a standard deviation scale. The association of genetically-determined NAFLD (first with 7 SNPs and then with 12 SNPs) and the blood metabolome was assessed using the IVW-MR with the *mr* function from *TwoSampleMR* package in R.^7^ The IVW-MR is comparable to performing a meta-analysis of each Wald ratio (the effect of the genetic instrument on the outcome divided by its effect on the exposure). Additional MR analysis were performed to evaluate horizontal pleiotropy (intercept p-value from MR Egger^30^) and the presence of outliers. We used MR-PRESSO^31^, an outlier-robust method, to detect the presence of outliers (variants potentially causing pleiotropy and influencing causal estimates) and causal estimates were obtained before and after excluding outliers. We also used, as sensitivity analyses, the simple median and weighted median consensus methods, which give unbiased causal inference if most genetic instruments are valid. Altogether, consistent results across different robust MR methods and significant p-value after correction for multiple testing give support to the robustness and provide further confirmation about the nature of the causal finding.

### Impact of NAFLD on tyrosine levels in the Estonian Biobank

Blood plasma levels of tyrosine were measured using nuclear magnetic resonance spectroscopy in 10,809 participants of the Estonian Biobank. Odds-ratios and corresponding p-values were estimated using logistic regression model implemented in R version 4.0.4^31^. Metabolite values were scaled and centered prior to analysis. Two models were run: raw model with adjusting for age and sex; and adjusted model, which was additionally adjusted for smoking status, education level and body-mass index. This study was approved by the Research Ethics Committee of the University of Tartu (Approval number 288/M-18).

### Impact of NAFLD and NASH on tyrosine levels in Québec bariatric surgery cohort

Plasma tyrosine levels were measured in 138 participants of the IUCPQ Obesity Biobank according to institutionally-approved management modalities. All participants provided written, informed consent. In this study sample, histological lesions from liver biopsy have been graded and staged using the protocol of Brunt at al.^32^ by pathologists who were blinded to the study objectives. We separated the participants in three groups based on the following three categories: 1) absence of hepatic steatosis (n=30), 2) hepatic steatosis without NASH (n=39) and 3) hepatic steatosis with NASH (n=69). Supplementary Table 4 presents the clinical characteristics at the time of surgery (sex, age, anthropometry, medication use, as well as glycaemic, lipoprotein and liver enzyme profile) for patients of each group (without NAFLD, NAFLD without NASH and NAFLD with NASH). From the plasma samples, a Water Acquity UPLC system coupled to a Synapt G2-Si mass spectrometer (Waters) in tandem mode (LC-MS/MS) using the EZ:faast amino acid sample testing kit (Phenomenex, 2003) was performed for the quantification of tyrosine levels. With this kit, plasma samples are mixed with an internal standard solution and amino acids are extract by a solid-phase support. Once extracted, amino acids are derivatized to increase stability for the analysis and purified by a two-phase liquid-liquid extraction. The samples were analyzed with the HPLC column, method gradient, and multiple monitoring (MRM) provided by the kit. For the last step of the quantification, a sample to internal standard ratios and a 5-point calibration curve ranging from 20 nmol/mL to 200 nmol/mL were used. An analysis of variance (ANOVA) followed by Tukey HSD test were performed to compare mean tyrosine levels between the three groups.

## Data Availability

All data produced in the present study are available upon reasonable request to the authors.

## Acknowledgements

We would like to thank all study participants as well as all investigators of the studies that were used throughout the course of this investigation. The Authors acknowledge the invaluable collaboration of the surgery team, bariatric surgeons, and biobank staff of the IUCPQ.

## Authors contributions

Conceptualization, B.J.A.; Methodology and Formal analysis, É.G., I.M-P., N.T., C.C; Writing—original draft preparation, É.G. and B.J.A.; Writing—review and editing, E.G., J.B., N.P., P.L.M., H.D.M.; Supervision, B.J.A. All authors have read and agreed to the published version of the manuscript.

